# Persistent Variations of Blood DNA Methylation Associated with Treatment Exposures and Risk for Cardiometabolic Outcomes among Long-term Survivors of Childhood Cancer: A Report from the St. Jude Lifetime Cohort

**DOI:** 10.1101/2020.09.10.20192393

**Authors:** Nan Song, Chia-Wei Hsu, Haitao Pan, Yinan Zheng, Lifang Hou, Jin-ah Sim, Zhenghong Li, Heather Mulder, John Easton, Emily Walker, Geoffrey Neale, Carmen L. Wilson, Kirsten K. Ness, Kevin R. Krull, Deo Kumar Srivastava, Yutaka Yasui, Jinghui Zhang, Melissa M. Hudson, Leslie L. Robison, I-Chan Huang, Zhaoming Wang

## Abstract

**Background:** It is well-established that cancer treatment substantially increases risk of long-term adverse health outcomes among childhood cancer survivors. However, there is limited research on the underlying mechanisms. To elucidate the pathophysiology and a possible causal pathway from treatment exposures to cardiometabolic conditions, we conducted epigenome-wide association studies (EWAS) to identify DNA methylation (DNAm) sites associated with cancer treatment exposures and examined whether treatment-associated DNAm sites mediate associations between specific treatments and cardiometabolic conditions.

**Methods:** We included 2,052 survivors (median age 33.7 years) of European ancestry from the St. Jude Lifetime Cohort Study, a retrospective hospital-based study with prospective clinical follow-up. Cumulative doses of chemotherapy and region-specific radiation were abstracted from medical records. Seven cardiometabolic conditions were clinically assessed. DNAm profile was measured using MethylationEPIC BeadChip with blood-derived DNA.

**Results:** By performing multiple treatment-specific EWAS, we identified 2,894 5’-cytosine-phosphate-guanine-3′ (CpG) sites mapped to 1,583 gene/regions associated with one or more cancer treatments at epigenome-wide significance level (P < 9×10^−8^). Among the treatment-associated CpGs, 298 were associated with obesity, 85 with hypercholesterolemia, 41 with hypertriglyceridemia, and four with abnormal glucose metabolism (False-Discovery-Rate-Adjusted P<0.05). We observed full mediation by methylation at five independent CpGs for association between abdominal field radiotherapy (abdominal-RT) and risk of hypertriglyceridemia, nearly full mediation (99.5%) by methylation at nine CpGs for association between abdominal-RT and hypercholesterolemia, and partial mediation (42.1%) by methylation at two CpGs for association between abdominal-RT and abnormal glucose metabolism. In addition, six CpGs partially mediated the association between brain-RT and obesity with 58.6% mediation effect, two CpGs mediated the association between glucocorticoids and obesity (32.2%) and between brain-RT and hypertriglyceridemia (15.5%). Notably, several mediator CpGs reside in the proximity of well-established dyslipidemia genes: cg17058475 (CPT1A), cg11851174 (RAI1) and cg22976567 (LMNA).

**Conclusions:** In childhood cancer survivors, prior cancer treatments are associated with DNAm variations present decades following the exposure. Treatment-associated DNAm sites may mediate the causal pathway from specific treatment exposures to certain cardiometabolic conditions, suggesting the utility of DNAm sites as risk predictors and potential mechanistic targets for future intervention studies.

## Background

Progress in cancer treatment has dramatically improved five-year survival following a childhood cancer diagnosis to more than 85% between 2010 and 2016.[1] Thus, the population of childhood cancer survivors has grown rapidly and is estimated to exceed 500,000 persons in the United States.[2] Unfortunately, the treatment of childhood malignancies is associated with long-term morbidity and mortality.[3–7] Mounting evidence suggests that reduced physical activity, muscular weakness, metabolic derangements, cognitive declines are common problems among adults treated for childhood malignancies.[7–11] Furthermore, premature cellular senescence, sterile inflammation, and mitochondrial dysfunction resulting from primary cancer diagnosis or treatment-related toxicity may contribute to adverse health outcomes.[12] Accordingly, survivors of childhood cancer often develop treatment-related late effects with 60% to more than 90% of survivors experiencing one or more long-term chronic health conditions (CHCs),[13] an approximately 2-fold greater burden of CHCs than community-controls.[4] Treatment-related adverse health outcomes encompass a broad range of CHCs,[4, 14, 15] hospitalizations,[16] premature frailty,[17] and early mortality.[14] Some of the most commonly observed CHCs among survivors include obesity,[18, 19] diabetes mellitus,[20] cardiovascular diseases,[21, 22] hypertension,[9, 22, 23] and subsequent neoplasms.[24, 25]

While the substantially increased risk and total burden of adverse health outcomes among childhood cancer survivors has been extensively described, there is a need to unravel the complex interplay between therapeutic exposures and genetic susceptibility in order to elucidate the pathogenesis of specific health conditions.[6] Pathogenic germline mutations in DNA repair genes contribute to subsequent neoplasm risk in childhood cancer survivors, especially among those who received high cumulative doses of specific agents and modalities.[26] Unlike germline genetics (DNA sequence), which is largely static throughout the life-course, epigenetic patterns are plastic and can be modified in response to internal and external insults including medical treatments.[27] Population-based studies among breast cancer[28] and gastric cancer[29]patients have provided evidence supporting that chemotherapy, radiotherapy, or a combination of anticancer treatments have a profound impact on epigenetic alterations, primarily in the form of CpG methylation. The processes leading to aberrant DNA methylation (DNAm) are poorly understood. How epigenetic alterations resulting from cancer therapy interact with downstream gene regulation machineries and ultimately lead to development of CHCs in individual survivors is still largely unknown. Possible biological mechanisms have been suggested; for instance, cellular oxidative stress and DNA damage can induce aberrant DNAm by recruiting DNA methyltransferase complex.[30] Emerging evidence suggests that alterations in DNAm in blood can at least influence immune regulation[31] or blood lipids and metabolites.[32] Several population-based studies of adult-onset cancers have identified treatment-induced blood DNAm changes and associated these changes with health outcomes, specifically cognitive decline in breast cancer patients[28] and poor survival for colorectal cancer,[33] lung cancer,[34, 35] and ovarian cancer patients.[36, 37] Thus, epigenetic alterations due to cancer therapeutic agents may mediate or modify gene regulation potentially resulting in systemic changes contributing to the development of CHCs.

It is biologically plausible that treatments used during the active childhood cancer could leave epigenetic marks. Hence, we hypothesized that cancer treatment modalities cause aberrant hypo- or hyper-DNAm, which may affect the long-term risk of CHC among childhood cancer survivors. In this study, epigenome-wide association studies (EWAS) were conducted among adult survivors of childhood cancer participating in the St. Jude Lifetime Cohort Study (SJLIFE) to identify differentially DNAm CpG sites between survivors exposed or unexposed to certain treatment and their associations with CHCs. We specifically focused on seven common cardiometabolic conditions including obesity, hypertension, hypercholesterolemia, hypertriglyceridemia, abnormal glucose metabolism, cardiomyopathy, and myocardial infarction, given that blood DNAm plays a role in the regulation of blood lipids and other metabolites.[32]

## Methods

### Study population

SJLIFE is a retrospective cohort study with prospective follow-up of survivors diagnosed with childhood cancer and treated at St. Jude Children’s Research Hospital, described elsewhere.[38, 39] Participants complete questionnaires assessing demographic and epidemiological factors and receive comprehensive medical and laboratory assessments at each follow-up visit to characterize their health conditions. Genome-wide EPIC methylation profiling (Illumina, San Diego, CA, USA) was performed using blood derived DNA from 2,689 SJLIFE survivors. Subsequent sample exclusion criteria included the following: 1) low total intensity of DNAm (n=3); 2) no whole-genome sequencing data (n=46); 3) age at blood draw under 18 years old (n=218); and 4) population admixture coefficient for CEU population <80% (n=370) based on the STRUCTURE analysis[40] with three continental references (JPT+CHB, CEU, YRI) from 1000 Genomes Project. Accordingly, we included 2,052 childhood cancer survivors of European ancestry in statistical analyses (Additional file 1: Figure S1).

### Treatment exposures

Treatment exposure information was extracted from medical records using a structured protocol.[38] Briefly, using radiation oncology treatment records, region-specific radiotherapy (RT) dosimetry, including brain-RT, chest-RT, abdominal-RT and pelvic-RT, was estimated.[41] Cumulative doses of individual chemotherapeutic agents including alkylating agents, anthracyclines, epipodophyllotoxins, glucocorticoids, and vincristine were abstracted from medical records. Equivalency approaches were applied for cumulative alkylating agents exposure[42] and anthracyclines exposure.[43]

### Chronic health conditions

A modification of the Common Terminology Criteria for Adverse Events (version 4.03, National Cancer Institute)[44] was applied to clinically ascertain medical outcomes and score for severity[39]. Clinical outcomes were severity-graded as 0 (no problem), 1 (mild), 2 (moderate), 3 (severe or disabling) and 4 (life-threatening)[39]. All CHCs with grades ≥1 were grouped together as cases. Considering DNAm is known to play an essential role in the regulation of blood lipids or metabolites[45], we included in this study seven common cardiometabolic CHCs: abnormal glucose metabolism, cardiomyopathy, hypercholesterolemia, hypertriglyceridemia, hypertension, myocardial infarction, and obesity. Only incident CHCs that occurred after blood draw for DNAm profiling were considered in the current study.

### DNA Methylation Profiling

Genome-wide DNA methylation data were generated using Infinium MethylationEPIC BeadChip array (Illumina, San Diego, CA, USA). Genomic DNA (250 ng) was extracted from blood samples according to standard procedures as described previously.[46] Further bisulfite treatment, array hybridization and scanning are provided in Supplementary Methods in the Additional file 1. The raw intensity data were exported from Illumina Genome Studio and analyzed in R (version 3.6.3) using minfi package[47]. Methylation level for a CpG site is described as a β value, which is a continuous variable ranging between 0 (no methylation) and 1 (full methylation). In any sample, a probe with a detection *P* value more than 0.01 was assigned missing status. Any sample or probe with more than 5% missing values were excluded from downstream analysis. Non-specific or cross-reactive probes, probes with SNPs nearby the CpG site or probes on sex chromosomes (X, Y) were also excluded. A total number of 686,880 probes remained for further analyses. Marker intensities were normalized by quantile normalization. M-value (i.e., logit transformation of β value) were subsequently calculated and used in regression analyses.[48] Six lymphocyte cell subtype proportions (lymphocyte, monocyte, and granulocyte, CD4, natural killer, and B cells) were estimated based on methylation signatures using Houseman’s method.[49, 50] A principal components analysis of methylation levels of all CpG sites that passed QC was performed to quantify latent structures or batch effects in the data. The array annotations provided by Illumina were used to map probes to their corresponding genes.

### Statistical analysis

To identify DNAm level at each CpG site influenced by specific treatment for childhood cancer, EWAS analysis was performed using multiple linear regression of methylation level at each CpG-site (dependent variable, continuous) on each treatment exposure status (independent variable, binary: exposed vs non-exposed) or the cumulative dose (independent variable, categorical, by tertiles or various dose ranges) with covariate adjustments including sex, age, other cancer treatment exposures (see below), lymphocyte subtype proportions, top three genetic principal components, and top four methylation principal components determined by the change rate of eigenvalues. Correlation between every pair of binary treatment exposures was described by Phi coefficient and the statistical significance (p-value) of its departure from 0 was assessed. Adjustment for other cancer treatment exposures in the EWAS was made if the Phi coefficient with the treatment exposure of interest is less than 0.4 and with a p-value > 0.05. R package CpGassoc[51] was used for the EWAS multiple linear regression analysis and we used *P* < 9×10^−8^ corresponding to 5% family-wise error as the threshold for genome-wide significance.[52] Manhattan plots were generated for visualization of EWAS results using CMplot R package[53] and Venn diagrams for visualizing unique and overlapping CpGs associated with different cancer treatments were generated using VennDiagram R package.[54] We further investigated associations between the residual methylation level of each treatment-associated methylation site (independent variable, residuals derived from the multiple linear regression of the EWAS model above but without adjusting for treatments) and a specific incident CHC (dependent variable, binary) using a multivariable logistic regression model adjusted for age and sex. False-discovery-rate (FDR)-adjusted p-values (P_*fdr*_) were obtained to control for multiple testing.[55] Survivors with specific CHC occurring prior to the collection of blood for DNAm profiling were excluded from this analysis. Mediation analysis was performed by treating CpG sites as hypothesized mediators for the association of treatment exposures with risk of incident CHCs using Mediation R package[56]. Statistical analysis workflow was summarized in Figure 2A. All statistical analyses were performed by using R.3.6.3[2] or SAS 9.4 (SAS Institute Inc., Cary, NC, USA) and all statistical tests were two-sided.

**Figure 2.**
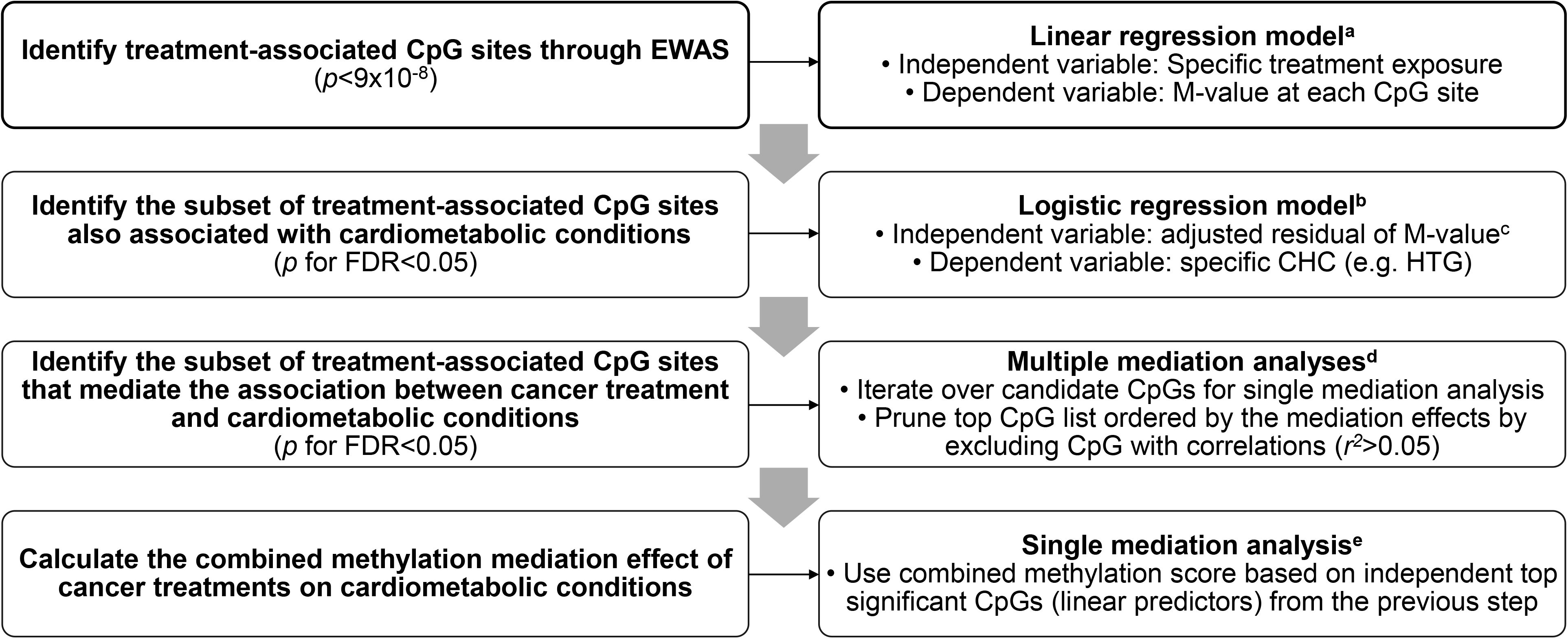
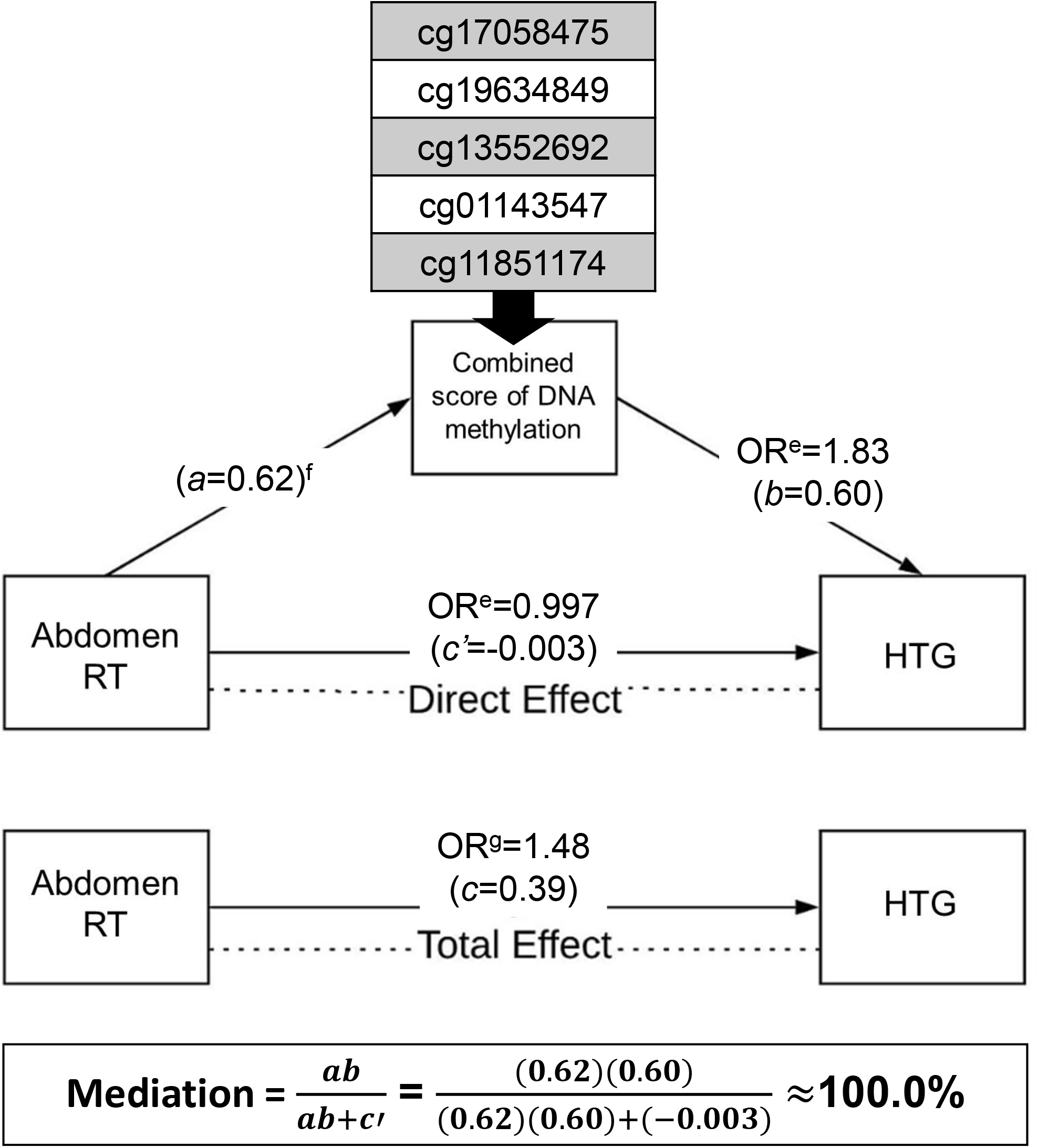
DNA methylation mediates the associations between treatment exposures and CHCs. (A) Statistical analysis workflow (B) Five DNA methylation CpG sites collectively mediate the effect of abdominal-RT on HTG risk Abbreviations: CHC (chronic health condition), EWAS (epigenome-wide association study), HTG (hypertriglyceridemia), RT (radiation therapy) ^a^Linear regression model was adjusted for covariates including sex, age, other cancer treatment exposures, lymphocyte subtype proportions, top three genetic principal components, and top four methylation principal components. ^b^logistic regression model was adjusted for covariates including sex and age^c^Residual of M-value was calculated based on linear regression adjusted for covariates including sex, age, lymphocyte subtype proportions, significant genetic principal components, and methylation principal components. ^d^Mediation analysis included two regression modes: a logistic regression model with CHC status as outcome, specific treatment as treatment variable (Term used for the exposure in Mediation R package), residual M-value for a CpG site as a mediator variable and adjusted for age, sex and other cancer treatment exposures; a linear regression model with residual M-value for a CpG site as outcome, specific treatment as treatment variable and other significant treatments as covariates. ^e^Mediation analysis as above except for replacing residual M-value for a CpG site with a combined methylation score by summing up the residual M-values for multiple CpG sites that were found to be significant mediators individually. ^f^a linear regression model with residual M-value for a CpG site as outcome, specific treatment as treatment variable and other significant treatments as covariates ^g^a logistic regression model with CHC status as outcome, a specific treatment as independent variable and adjusted for sex, age and other significant treatments as covariates

## Results

### Characteristics of study population

Table 1 shows the characteristics of the 2,052 childhood cancer survivors of European ancestry. Survivors were previously diagnosed with leukemia (34.1%), lymphoma (21.8%), sarcoma (13.4%), central nervous system (CNS) tumors (11.3%), embryonal tumors (13.5%), and others (6.0%). Treatment exposures comprised alkylating agents (58.2%), anthracyclines (58.0%), epipodophyllotoxins (34.6%), glucocorticoids (47.0%), vincristine (69.2%), brain-RT (30.7%), chest-RT (28.1%), abdominal-RT (20.1%), and pelvic-RT (17.2%). The incidence of cardiometabolic conditions in the study population was: abnormal glucose metabolism (18.0%, 95% CI=16.2-19.9%), cardiomyopathy (9.6%, 95% CI=8.4-11.1%), hypercholesterolemia (32.8%, 95% CI=30.5-35.3%), hypertriglyceridemia (25.9%, 95% CI=23.8-28.2%), hypertension (53.3%, 95% CI=50.7-56.0%), myocardial infarction (2.5%, 95% CI=1.9-3.3%), and obesity (62.0%, 95% CI=59.6-64.5%). The median age at diagnosis was 8.5 (range=0.0-23.6) years and the median age at DNA sampling was 33.7 (range=18.0-66.4) years.

**Table 1.** Characteristics of study population

| Characteristics | N | (%) |
| --- | --- | --- |
| Total | 2,052 | (100.0) |
| Sex |  |  |
| Male | 1,084 | (52.8) |
| Female | 968 | (47.2) |
| Diagnosis |  |  |
| Leukemia | 699 | (34.1) |
| Acute lymphoblastic leukemia | 644 | (31.4) |
| Acute myeloid leukemia | 53 | (2.6) |
| Other leukemia | 2 | (0.1) |
| Lymphoma | 448 | (21.8) |
| Hodgkin lymphoma | 288 | (14.0) |
| Non-Hodgkin lymphoma | 160 | (7.8) |
| Sarcoma | 274 | (13.4) |
| Ewing sarcoma | 74 | (3.6) |
| Osteosarcoma | 74 | (3.6) |
| Rhabdomyosarcoma | 71 | (3.5) |
| Soft tissue sarcoma | 55 | (2.7) |
| CNS tumors | 231 | (11.3) |
| Astrocytoma or glioma | 93 | (4.5) |
| Medulloblastoma or PNET | 56 | (2.7) |
| Ependymoma | 26 | (1.3) |
| Other CNS tumors | 56 | (2.7) |
| Embryonal | 276 | (13.5) |
| Wilms tumor | 134 | (6.5) |
| Neuroblastoma | 107 | (5.2) |
| Germ cell tumor | 35 | (1.7) |
| Other | 124 | (6.0) |
| Retinoblastoma | 45 | (2.2) |
| Hepatoblastoma | 13 | (0.6) |
| Melanoma | 12 | (0.6) |
| Carcinomas | 24 | (1.2) |
| Others | 30 | (1.5) |
| Chemotherapy |  |  |
| Alkylating agent | 1,194 | (58.2) |
| Anthracyclines | 1,190 | (58.0) |
| Epipodophyllotoxins | 709 | (34.6) |
| Glucocorticoids | 965 | (47.0) |
| Vincristine | 1,420 | (69.2) |
| Radiation therapy (RT) |  |  |
| Brain-RT | 629 | (30.7) |
| Chest-RT | 577 | (28.1) |
| Abdominal-RT | 412 | (20.1) |
| Pelvic-RT | 352 | (17.2) |
| CHCs | Incident N/N at risk | (%, 95% CI) |
| Abnormal glucose metabolism | 302/1,680 | (18.0, 16.2-19.9) |
| Cardiomyopathy | 168/1,742 | (9.6, 8.4-11.1) |
| Hypercholesterolemia | 479/1,461 | (32.8, 30.5-35.3) |
| Hypertriglyceridemia | 399/1,539 | (25.9, 23.8-28.2) |
| Hypertension | 727/1,365 | (53.3, 50.7-56.0) |
| Myocardial infarction | 47/1,892 | (2.5, 1.9-3.3) |
| Obesity | 942/1,519 | (62.0, 59.6-64.5) |
| Median age at diagnosis, years (range) | 8.5 | (0.0-23.6) |
| Median age at DNA sampling, years (range) | 33.7 | (18.0-66.4) |

Abbreviations: CNS (central nervous system), PNET (primitive neuroectodermal tumor), RT (radiation therapy), CHC (chronic health condition), and CI (confidence interval)

### Treatment-specific associations of DNA methylation

Examination of all the pairwise correlations among the nine different treatments (Additional file 1: Table S1) found moderate to high correlations between alkylating agents and vincristine, corticosteroids and vincristine, chest-RT and abdominal-RT, and chest-RT and pelvic-RT (Phi coefficient > 0.4 and *P* < 0.05). By performing multiple treatment-specific EWAS analyses after excluding the strongly-correlated treatments from covariate adjustments, a total of 2,894 CpG sites mapped to 1,583 gene/regions were associated with one or more cancer treatments at epigenome-wide significant level (P < 9×10^−8^). These epigenome-wide results showed 1,028 DNAm hits for alkylating agents, 548 hits for epipodophyllotoxin, 22 hits for glucocorticoids, 36 hit for vincristine, 69 hits for brain-RT, 1,388 hits for chest-RT, 1,495 hits for abdominal-RT, 895 hits for pelvic-RT, but none for anthracyclines (Additional file 1: Figure S2). Figure 1 shows the overlap of DNAm sites associated with specific chemotherapeutic agent or RT field. Among 1,353 CpG sites identified to be associated with chemotherapy exposure, there were 255 CpG sites associated with two or more chemotherapy agents and the remaining were specifically associated with alkylating agents (n=783), epipodophyllotoxins (n=304), glucocorticoids (n=6), and vincristine (n=5) (Figure 1A). Among 2,094 CpG sites associated with RT exposure, there were 794 CpG sites associated with two or more region-specific RT treatments and the remaining are specifically associated with brain-RT (n=68), chest-RT (n=409), abdominal-RT (n=366), or pelvic-RT (n=102) (Figure 1B).

**Figure 1.**
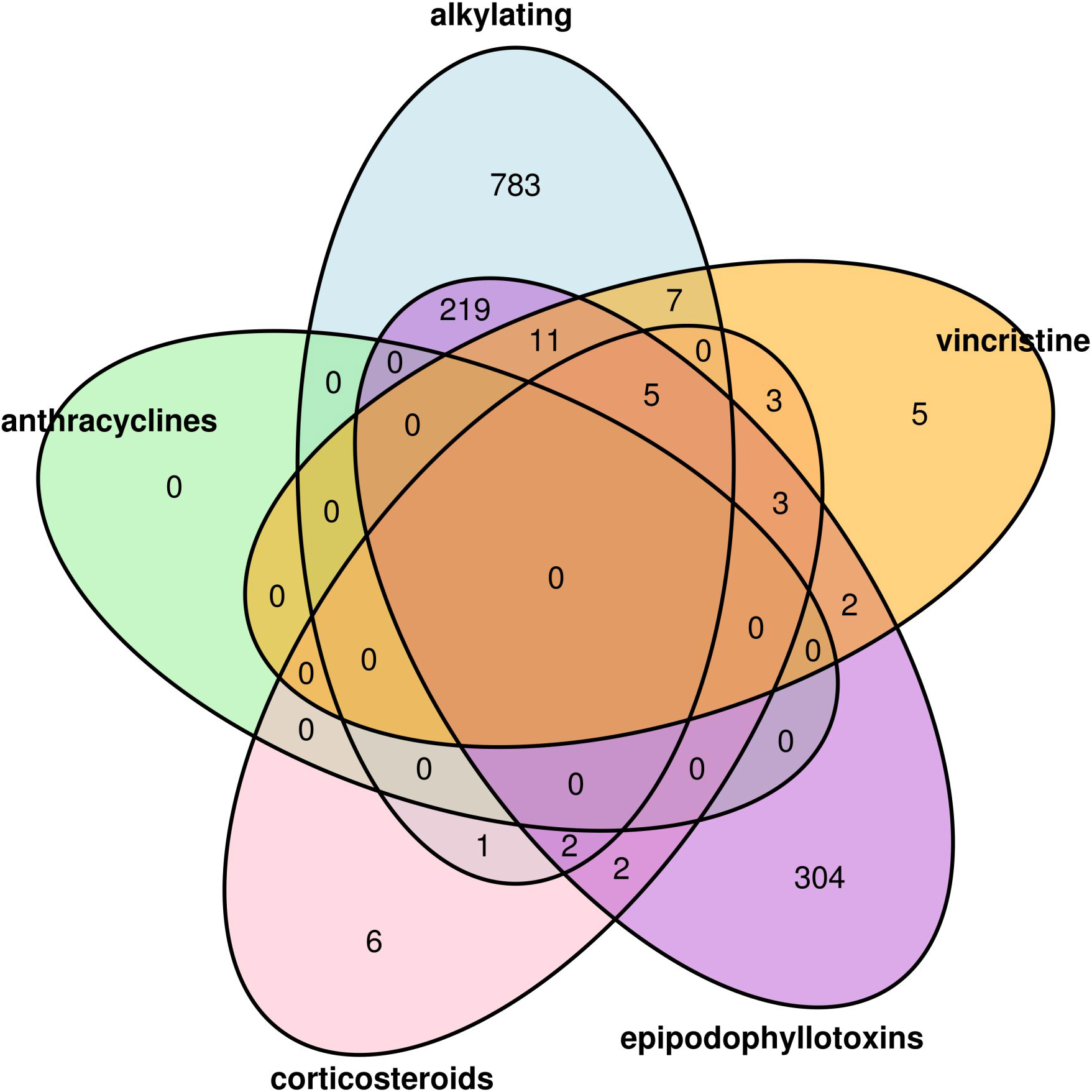
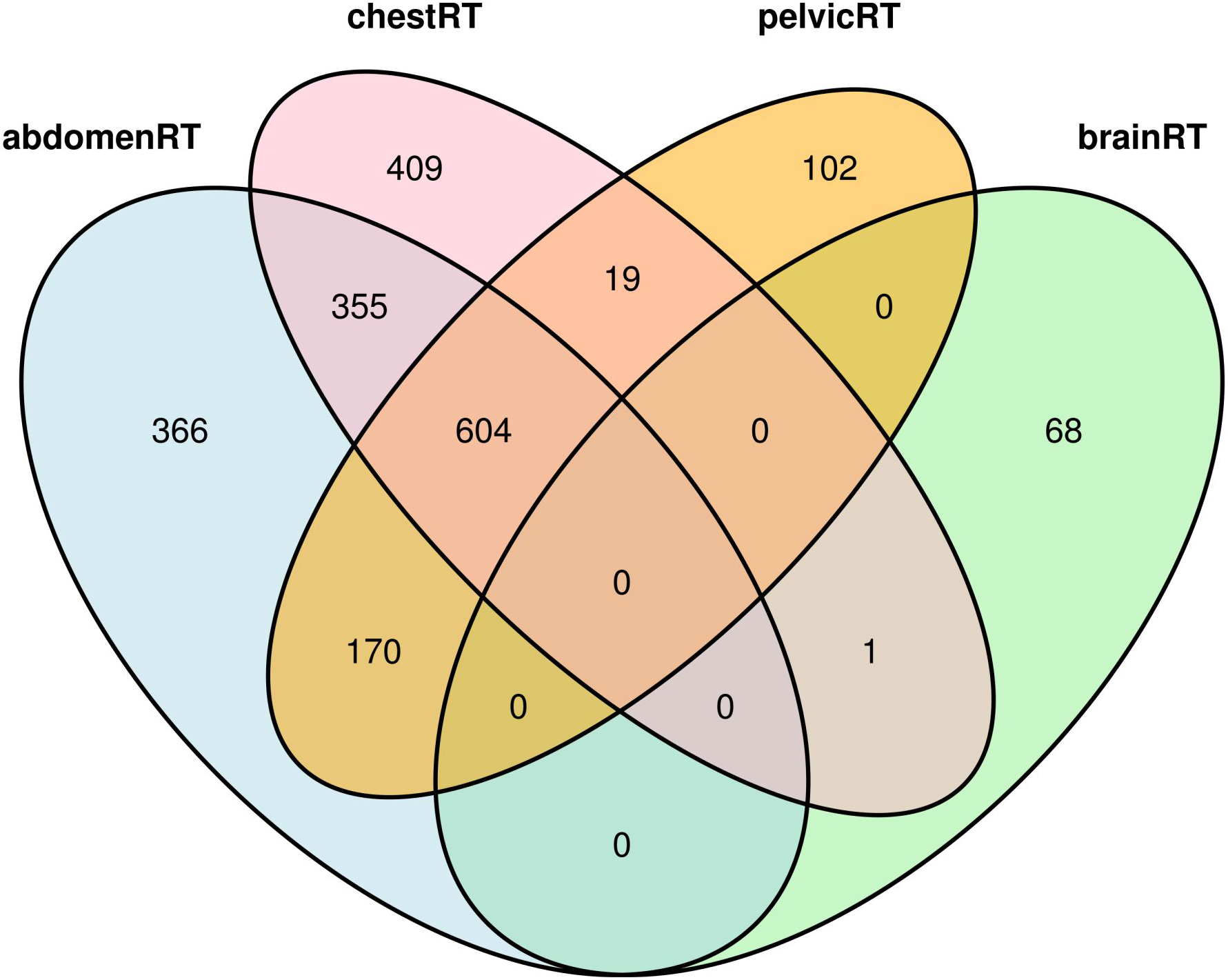
Venn Diagram showing the overlap of DNA methylation sites associated with specific cancer treatments. (A) Chemotherapy (B) Radiation therapy

Among significant CpGs for each treatment status, there were also statistically significant linear dose-response relationship between the continuous cumulative dose of the specific treatment and DNAm level showing 13/1,028 (1.3%) hits for alkylating agents, 8/22 (36.4%) hits for glucocorticoids, 110/548 (20.1%) hits for epipodophyllotoxins, 7/36 (19.4%) hits for vincristine, 60/69 (87.0%) hits for brain-RT, 1,267/1,388 (91.3%) hits for chest-RT, 1,157/1,495 (77.4%) hits for abdominal-RT, and 777/895 (86.8%) hits for pelvic-RT.

### Association of treatment-associated methylation sites with CHC

Evaluation of the association between each treatment-related methylation site (n=2,894) and each CHC using multivariable logistic regression models found the highest number of CpGs were significantly (P_fdr_ < 0.05) associated with obesity (n=298), followed by hypercholesterolemia (n=85), hypertriglyceridemia (n=41) and abnormal glucose metabolism (n=4) (Additional file 1: Table S2). There was no single CpG site associated with cardiomyopathy, myocardial infarction or hypertension.

### Treatment-associated methylation sites mediate effect of treatment on CHC

The associations between treatment exposures and CHCs are presented in Additional file 1: Table S3. Abdominal-RT was found to be associated with increased risk of hypertriglyceridemia with an odds ratio (OR) of 1.48 (95% CI = 1.11-1.98). There were 29 CpG sites whose methylation levels were associated with abdominal-RT at genome-wide significance level and also with hypertriglyceridemia after adjusting for multiple comparison. In the mediation analysis, each of these 29 CpGs was considered as a hypothesized mediator variable, abdominal-RT as the exposure and status of hypertriglyceridemia (binary) as the outcome while adjusting for sex, age and brain-RT (another exposure significantly associated with hypertriglyceridemia). Twenty-four CpGs were identified with significant average causal mediation effects (ACME) (P_fdr_ < 0.05). Using pairwise Pearson correlation coefficient r^2^ threshold of 0.05, five independent CpGs were obtained by top-down pruning of the 24 CpGs sorted by estimated ACME in decreasing order. For the final mediation analysis, using a combined score (i.e., summation of the methylation levels of five CpGs) as a mediator variable, full mediation (100%) was achieved (Figure 2B and Table 2). Using the same strategy, we found a set of two CpGs partially mediated (42%) the effect of abdominal-RT on increased risk of abnormal glucose metabolism (OR= 1.57, 95% CI=1.16-2.12), a combined score encompassing 9 CpGs accounted for 99.5% mediated effects of abdominal-RT on risk of hypercholesterolemia (OR= 1.54, 95% CI=1.15-2.06), and another set of two CpGs partially mediated (32.2%) the effect of corticosteroids on risk of obesity (OR= 1.56, 95% CI=1.24-1.95).

**Table 2.** DNA methylation CpG sites mediate the associations between treatment exposures and CHCs

| Treatment | CHC | Total Effect, OR | CpG | Chromosome | Gene | % Mediation |
| --- | --- | --- | --- | --- | --- | --- |
| Abdominal-RT | HTG | 1.48 | cg17058475 | chr11 | *CPT1A* | 41.3 |
|  |  |  | cg19634849 | chr5 | *CYSTM1;PFDN1* | 29.2 |
|  |  |  | cg13552692 | chr18 | *CCDC102B* | 26.0 |
|  |  |  | cg01143547 | chr6 | *CCND3* | 16.7 |
|  |  |  | cg11851174 | chr17 | *RAI1* | 14.8 |
|  |  |  | Combined score<sup>a</sup> |  |  | 100.0 |
| Brain-RT | HTG | 1.71 | cg01358620 | chr14 | *HEATR5A;RP11-176H8.1* | 9.7 |
|  |  |  | cg24018185 | chr12 | NA (gene desert) | 8.4 |
|  |  |  | Combined score<sup>a</sup> |  |  | 15.5 |
| Abdominal-RT | HCL | 1.54 | cg01511232 | chr4 | *LRAT* | 37.8 |
|  |  |  | cg22976567 | chr1 | *LMNA* | 20.0 |
|  |  |  | cg17058475 | chr11 | *CPT1A* | 19.6 |
|  |  |  | cg12939283 | chr6 | *HLA-DPA1;HLA-DPB1* | 17.9 |
|  |  |  | cg07398622 | chr6 | NA | 16.5 |
|  |  |  | cg12956751 | chr2 | *AC068134.8;ALPP* | 13.4 |
|  |  |  | cg13562176 | chr16 | *HMOX2;NMRAL1* | 13.1 |
|  |  |  | cg06917935 | chr8 | *ZFPM2;ZFPM2-AS1* | 13.0 |
|  |  |  | cg10562405 | chr17 | *LINC00511* | 11.4 |
|  |  |  | Combined score<sup>a</sup> |  |  | 99.5 |
| Abdominal-RT | AGM | 1.57 | cg14263063 | chr16 | *DNAH3* | 30.1 |
|  |  |  | cg09006543 | chr13 | *FRY* | 14.6 |
|  |  |  | Combined score<sup>a</sup> |  |  | 42.0 |
| Brain-RT | Obesity | 1.57 | cg12098187 | chr10 | *LCOR* | 20.9 |
|  |  |  | cg09174411 | chr12 | *BTBD11* | 18.2 |
|  |  |  | cg26572901 | chr7 | NA | 14.9 |
|  |  |  | cg17739917 | chr17 | *RARA* | 14.3 |
|  |  |  | cg12715065 | chr6 | NA | 13.3 |
|  |  |  | cg04173586 | chr19 | *DOT1L* | 11.8 |
|  |  |  | Combined score<sup>a</sup> |  |  | 58.6 |
| Glucocorticoids | Obesity | 1.56 | cg22351187 | chr12 | *KRT80* | 19.0 |
|  |  |  | cg03684356 | chr20 | *PMEPA1* | 16.3 |
|  |  |  | Combined score<sup>a</sup> |  |  | 32.2 |

Abbreviations: CHC (chronic health condition), OR (odds ratio), RT (radiation therapy), HTG (hypertriglyceridemia), HCL (hypercholesterolemia), AGM (abnormal glucose metabolism)
<sup>a</sup>Calculated by summing DNA methylation values of CpG sites with individually significant mediation effect of treatment exposures and CHCs.

## Discussion

The biological basis underlying treatment-related risks for adverse health outcomes among survivors of childhood cancer is largely unknown. We speculated that one plausible casual pathway is the acquisition and persistent soma-wide alterations in DNAm. In this study, the first large-scale association analyses between cancer treatments and DNA methylation in survivors of childhood cancer, our mediation analyses provide compelling evidence in substantiating this hypothesis. Moreover, we identify unique and overlapping DNAm signatures across different cancer treatments that may serve as mechanistic targets for future intervention studies.

Many of the genome-wide significant treatment-associated CpGs have established associations with aging, smoking, diet and other lifestyle factors[57], suggesting there is a common set of CpGs serving as “sensors” that are sensitive to both internal and external environments. Among chemotherapy exposures, alkylating agents had the highest number of CpG hits, followed by epipodophyllotoxins, vincristine and glucocorticoids, and there was no single hit for anthracyclines. The majority of alkylating agent associated CpGs (65%) were demethylated, which is consistent with an oxidative demethylation process as one mechanism to repair DNA alkylation damage.[58, 59] Among radiation exposures, there were a comparable number of hits among chest-RT, abdominal-RT, and pelvic-RT exposures, but far fewer CpGs hits associated with brain-RT exposure. This is likely due to limited exposure to the bone marrow tissue among patients who received RT to the brain, considering we measured methylation on blood derived DNA. In this regard, the observation of a high percentage of CpGs with a dose response for RT exposures, but not for chemotherapy agents, was intriguing.

Some of the sensor CpGs were also associated with human traits such as aging, dyslipidemia, BMI and alcohol consumption based on the annotation using EWAS catalog (Additional file 1: Table S4) [57] as well as one or more of the seven CHCs in our study, which make them eligible as potential mediators in mediation analysis for the pathway from treatment exposures to health outcomes. Indeed, we found a range of mediation effects by multiple CpGs for associations between treatments and CHCs. It is highly notable that we found 100% mediation for association between abdominal-RT and hypertriglyceridemia or hypercholesterolemia. The existing literature provides strong support for the plausibility of our findings. DNAm has an established role in the regulation of blood lipids and the etiology of dyslipidemia.[45] The mediator CpGs, that showed statistically significant mediation effects in this study have been associated with blood lipids and related diseases in other studies and/or are in the proximity of well-established genes regulating dyslipidemia. (e.g., cg17058475 (CPT1A),[60] cg11851174 (RAI1),[61] and cg22976567 (LMNA)).[62] The identification of key genes previously implicated in abnormal lipid metabolism in an agnostic EWAS attests to the strength of our findings.

For the partially mediated effects we discovered, other causal pathways from specific treatment to CHC are possible, including a process that indicates that DNA damage is associated with anticancer therapies and specific mechanisms for DNA repair.[63–67] Our previous study showed pathogenic germline variant in DNA repair pathways increase risk of developing subsequent neoplasms, especially among survivors who received high doses of radiation or specific types of chemotherapeutic agents.[26]

Our study has some limitations. First, due to frequent use of multimodality therapy, delineation of independent associations was not always feasible. To identify independent hits for each treatment, EWAS for each treatment was adjusted for other treatment exposures except for specific treatments that were highly correlated. Second, we did not consider other factors that affect the methylation landscape such as social economic status, health behaviors, and environmental exposures which could confound the findings. Third, even though we considered temporality among treatment exposures, DNAm (measured at a single time point), and incidence of CHCs, we could not definitively infer causality amongst these three entities. Lastly, our study focused on survivors of European-ancestry. Further replication with larger and more diverse survivor populations and validation to confirm generalizability to other races/ethnicities are needed to confirm the role of DNAm in associations between cancer treatments and adverse health outcomes.

## Conclusions

In summary, we identified thousands of CpG sites associated with specific cancer treatments at genome-wide significant levels, suggesting that DNAm is an important biological embedding mechanism for prior cancer treatment exposures. We observed hundreds of these treatment-associated CpG sites significantly associated with one or more of the seven cardiometabolic CHC risk after adjusting for multiple testing. Moreover, dozens of these sensor CpG sites showed full or partial mediation effects for the association between specific treatment exposure and cardiometabolic CHC, suggesting DNAm, as a biomarker, can be used as a risk predictor and potential mechanistic target for future intervention studies to mitigate long-term treatment toxicity among survivors of childhood cancer.

## Data Availability

The St. Jude Lifetime Cohort Study data are accessible through the St. Jude Cloud (https://stjude.cloud).

## List of abbreviations

ACME: average causal mediation effects
AGM: abnormal glucose metabolism
CHC: chronic health conditions
CI: confidence interval
CNS: central nervous system
CpG: 5’-cytosine phosphate-guanine-3′
EWAS: epigenome-wide association study
FDR: false-discovery-rate
HCL: hypercholesterolemia
HTG: hypertriglyceridemia
OR: odds ratio
PNET: primitive neuroectodermal tumor
RT: radiotherapy
SJLIFE: St. Jude Lifetime Cohort Study

## Declarations

### Ethics approval and consent to participate

All SJLIFE study participants provided written informed consent. The SJLIFE study protocol was approved by the Institutional Review Board (IRB) at SJCRH.

## Consent for publication

Not applicable

## Availability of data and materials

The St. Jude Lifetime Cohort Study data are accessible through the St. Jude Cloud (https://stjude.cloud). Z.W. had full access to all the data in the study and takes responsibility for the integrity of the data and the accuracy of the data analysis.

## Authors’ contributions

ZW, I-CH, and LLR designed the study. MMH and LLR assisted in or provided support for data collection and recruitment of study participants. JE, HM, and CLW assisted in or supervised sample collection and processing; JE, HM, EP, GN, EW, and JZ supervised and/or performed DNA extraction and library preparation for the Infinium MethylationEPIC Array. NS and ZW performed the DNA methylation analysis. ZW, and HP developed the statistical analysis plan. CWH and NS performed the statistical analyses. NS, and ZW wrote the first draft of the manuscript. All authors contributed to data interpretation and writing and approved the final manuscript for publication.

## Additional files

**Additional file 1: Supplementary Methods**. Bisulfite treatment, array hybridization, and scanning for DNA Methylation Profiling. **Figure S1**.A flow diagram of the study population. **Figure S2**.Manhattan plot showing the treatment-specific association of DNA methylation. **Table S1**. Pairwise correlations among nine different treatments. **Table S2**. Association of treatment-associated methylation sites with cardiometabolic CHCs (P_FDR_< 0.05). **Table S3**. Multivariable associations of treatment exposures with cardiometabolic CHCs (P< 0.05). **Table S4**. Previously published methylation associations of CpG sites from blood derived DNA with health conditions.

## References

1. Siegel RL, Miller KD, Jemal A. Cancer statistics, 2019. CA Cancer J Clin. 2019;69(1):7–34.

2. Robison LL, Hudson MM. Survivors of childhood and adolescent cancer: life-long risks and responsibilities. Nature reviews Cancer. 2014;14(1):61–70.

3. Phillips SM, Padgett LS, Leisenring WM, Stratton KK, Bishop K, Krull KR, et al. Survivors of childhood cancer in the United States: prevalence and burden of morbidity. Cancer Epidemiol Biomarkers Prev. 2015;24(4):653–63.

4. Bhakta N, Liu Q, Ness KK, Baassiri M, Eissa H, Yeo F, et al. The cumulative burden of surviving childhood cancer: an initial report from the St Jude Lifetime Cohort Study (SJLIFE). Lancet (London, England). 2017;390(10112):2569–82.

5. Hudson MM, Ness KK, Gurney JG, Mulrooney DA, Chemaitilly W, Krull KR, et al. Clinical ascertainment of health outcomes among adults treated for childhood cancer. JAMA. 2013;309(22):2371–81.

6. Armenian SH, Bhatia S. Chronic health conditions in childhood cancer survivors: is it all treatment-related--or do genetics play a role? Journal of general internal medicine. 2009;24 Suppl 2(Suppl 2):S395–400.

7. Oeffinger KC, Mertens AC, Sklar CA, Kawashima T, Hudson MM, Meadows AT, et al. Chronic health conditions in adult survivors of childhood cancer. The New England journal of medicine. 2006;355(15):1572–82.

8. Meacham LR, Chow EJ, Ness KK, Kamdar KY, Chen Y, Yasui Y, et al. Cardiovascular risk factors in adult survivors of pediatric cancer--a report from the childhood cancer survivor study. Cancer Epidemiol Biomarkers Prev. 2010;19(1):170–81.

9. Oeffinger KC, Adams-Huet B, Victor RG, Church TS, Snell PG, Dunn AL, et al. Insulin resistance and risk factors for cardiovascular disease in young adult survivors of childhood acute lymphoblastic leukemia. Journal of clinical oncology: official journal of the American Society of Clinical Oncology. 2009;27(22):3698–704.

10. Janiszewski PM, Oeffinger KC, Church TS, Dunn AL, Eshelman DA, Victor RG, et al. Abdominal obesity, liver fat, and muscle composition in survivors of childhood acute lymphoblastic leukemia. The Journal of clinical endocrinology and metabolism. 2007;92(10):3816–21.

11. Tonorezos ES, Vega GL, Sklar CA, Chou JF, Moskowitz CS, Mo Q, et al. Adipokines, body fatness, and insulin resistance among survivors of childhood leukemia. Pediatric blood & cancer. 2012;58(1):31–6.

12. Ness KK, Armstrong GT, Kundu M, Wilson CL, Tchkonia T, Kirkland JL. Frailty in childhood cancer survivors. Cancer. 2015;121(10):1540–7.

13. Board PDQPTE. Late Effects of Treatment for Childhood Cancer (PDQ®): Health Professional Version. PDQ Cancer Information Summaries. Bethesda (MD): National Cancer Institute (US); 2002.

14. Armstrong GT, Chen Y, Yasui Y, Leisenring W, Gibson TM, Mertens AC, et al. Reduction in Late Mortality among 5-Year Survivors of Childhood Cancer. The New England journal of medicine. 2016;374(9):833–42.

15. Henson KE, Reulen RC, Winter DL, Bright CJ, Fidler MM, Frobisher C, et al. Cardiac Mortality Among 200 000 Five-Year Survivors of Cancer Diagnosed at 15 to 39 Years of Age:The Teenage and Young Adult Cancer Survivor Study. Circulation. 2016;134(20):1519–31.

16. Zhang Y, Lorenzi MF, Goddard K, Spinelli JJ, Gotay C, McBride ML. Late morbidity leading to hospitalization among 5-year survivors of young adult cancer: a report of the childhood, adolescent and young adult cancer survivors research program. International journal of cancer. 2014;134(5):1174–82.

17. Ness KK, Krull KR, Jones KE, Mulrooney DA, Armstrong GT, Green DM, et al. Physiologic frailty as a sign of accelerated aging among adult survivors of childhood cancer: a report from the St Jude Lifetime cohort study. Journal of clinical oncology: official journal of the American Society of Clinical Oncology. 2013;31(36):4496–503.

18. Garmey EG, Liu Q, Sklar CA, Meacham LR, Mertens AC, Stovall MA, et al. Longitudinal changes in obesity and body mass index among adult survivors of childhood acute lymphoblastic leukemia: a report from the Childhood Cancer Survivor Study. Journal of clinical oncology: official journal of the American Society of Clinical Oncology. 2008;26(28):4639–45.

19. Meacham LR, Gurney JG, Mertens AC, Ness KK, Sklar CA, Robison LL, et al. Body mass index in long-term adult survivors of childhood cancer: a report of the Childhood Cancer Survivor Study. Cancer. 2005;103(8):1730–9.

20. Meacham LR, Sklar CA, Li S, Liu Q, Gimpel N, Yasui Y, et al. Diabetes mellitus in long-term survivors of childhood cancer. Increased risk associated with radiation therapy: a report for the childhood cancer survivor study. Arch Intern Med. 2009;169(15):1381–8.

21. Bowers DC, McNeil DE, Liu Y, Yasui Y, Stovall M, Gurney JG, et al. Stroke as a late treatment effect of Hodgkin’s Disease: a report from the Childhood Cancer Survivor Study. Journal of clinical oncology: official journal of the American Society of Clinical Oncology. 2005;23(27):6508–15.

22. van der, Pal HJ, van Dalen EC, Kremer LC, Bakker PJ, van Leeuwen FE. Risk of morbidity and mortality from cardiovascular disease following radiotherapy for childhood cancer: a systematic review. Cancer Treat Rev. 2005;31(3):173–85.

23. Nuver J, Smit AJ, Postma A, Sleijfer DT, Gietema JA. The metabolic syndrome in long-term cancer survivors, an important target for secondary preventive measures. Cancer Treat Rev. 2002;28(4):195–214.

24. Armstrong GT, Liu W, Leisenring W, Yasui Y, Hammond S, Bhatia S, et al. Occurrence of multiple subsequent neoplasms in long-term survivors of childhood cancer: a report from the childhood cancer survivor study. Journal of clinical oncology: official journal of the American Society of Clinical Oncology. 2011;29(22):3056–64.

25. Meadows AT, Friedman DL, Neglia JP, Mertens AC, Donaldson SS, Stovall M, et al. Second neoplasms in survivors of childhood cancer: findings from the Childhood Cancer Survivor Study cohort. Journal of clinical oncology: official journal of the American Society of Clinical Oncology. 2009;27(14):2356–62.

26. Qin N, Wang Z, Liu Q, Song N, Wilson CL, Ehrhardt MJ, et al. Pathogenic Germline Mutations in DNA Repair Genes in Combination With Cancer Treatment Exposures and Risk of Subsequent Neoplasms Among Long-Term Survivors of Childhood Cancer. Journal of clinical oncology: official journal of the American Society of Clinical Oncology. 2020:Jco1902760.

27. Relton CL, Davey Smith G. Epigenetic epidemiology of common complex disease: prospects for prediction, prevention, and treatment. PLoS medicine. 2010;7(10):e1000356.

28. Yao S, Hu Q, Kerns S, Yan L, Onitilo AA, Misleh J, et al. Impact of chemotherapy for breast cancer on leukocyte DNA methylation landscape and cognitive function: a prospective study. Clin Epigenetics. 2019;11(1):45.

29. Choi SJ, Jung SW, Huh S, Chung YS, Cho H, Kang H. Alteration of DNA Methylation in Gastric Cancer with Chemotherapy. Journal of microbiology and biotechnology.2017;27(8):1367–78.

30. O’Hagan HM, Wang W, Sen S, Destefano Shields C, Lee SS, Zhang YW, et al. Oxidative damage targets complexes containing DNA methyltransferases,SIRT1, and polycomb members to promoter CpG Islands. Cancer Cell.2011 20(5):606–19.

31. Li H, Chiappinelli KB, Guzzetta AA, Easwaran H, Yen RW, Vatapalli R, et al. Immune regulation by low doses of the DNA methyltransferase inhibitor 5-azacitidine in common human epithelial cancers. Oncotarget. 2014;5(3):587–98.

32. Samblas M, Milagro FI, Martínez A. DNA methylation markers in obesity, metabolic syndrome, and weight loss. Epigenetics. 2019;14(5):421–44.

33. Fouad MA, Salem SE, Hussein MM,Zekri ARN, Hafez HF, El Desouky ED, et al. Impact of Global DNA Methylation in Treatment Outcome of Colorectal Cancer Patients. Frontiers in pharmacology.2018;9:1173.

34. Mo ML, Ma J, Chen Z, Wei B, Li H, Zhou Y, et al. Measurement of genome-wide DNA methylation predicts survival benefits from chemotherapy in non-small cell lung cancer. Journal of cancer research and clinical oncology. 2015;141(5):901–8.

35. Salazar F, Molina MA, Sanchez-Ronco M, Moran T, Ramirez JL, Sanchez JM, et al. First22 line therapy and methylation status of CHFR in serum influence outcome to chemotherapy versus EGFR tyrosine kinase inhibitors as second-line therapy in stage IV non-small-cell lung cancer patients. Lung cancer (Amsterdam, Netherlands). 2011;72(1):84–91.

36. Flanagan JM, Wilson A, Koo C, Masrour N, Gallon J, Loomis E, et al. Platinum-Based Chemotherapy Induces Methylation Changes in Blood DNA Associated with Overall Survival in Patients with Ovarian Cancer. Clin Cancer Res. 2017;23(9):2213–22.

37. Gifford G, Paul J, Vasey PA, Kaye SB, Brown R. The acquisition of hMLH1 methylation in plasma DNA after chemotherapy predicts poor survival for ovarian cancer patients. Clin Cancer Res. 2004;10(13):4420–6.

38. Hudson MM, Ness KK, Nolan VG, Armstrong GT, Green DM, Morris EB, et al. Prospective medical assessment of adults surviving childhood cancer: study design, cohort characteristics, and feasibility of the St. Jude Lifetime Cohort study. Pediatric blood & cancer. 2011;56(5):825–36.

39. Hudson MM, Ehrhardt MJ, Bhakta N, Baassiri M, Eissa H, Chemaitilly W, et al. Approach for Classification and Severity Grading of Long-term and Late-Onset Health Events among Childhood Cancer Survivors in the St. Jude Lifetime Cohort. Cancer Epidemiol Biomarkers Prev. 2017;26(5):666–74.

40. Porras-Hurtado L, Ruiz Y, Santos C, Phillips C, Carracedo A, Lareu MV. An overview of STRUCTURE: applications, parameter settings, and supporting software. Front Genet. 2013;4:98.

41. Stovall M, Donaldson SS, Weathers RE, Robison LL, Mertens AC, Winther JF, et al. Genetic effects of radiotherapy for childhood cancer: gonadal dose reconstruction. International journal of radiation oncology, biology, physics. 2004;60(2):542–52.

42. Green DM, Nolan VG, Goodman PJ, Whitton JA, Srivastava D, Leisenring WM, et al. The cyclophosphamide equivalent dose as an approach for quantifying alkylating agent exposure: a report from the Childhood Cancer Survivor Study. Pediatric blood & cancer. 2014;61(1):53–67.

43. Feijen EA, Leisenring WM, Stratton KL, Ness KK, van der Pal HJ, Caron HN, et al. Equivalence Ratio for Daunorubicin to Doxorubicin in Relation to Late Heart Failure in Survivors of Childhood Cancer. Journal of clinical oncology: official journal of the American Society of Clinical Oncology. 2015;33(32):3774–80.

44. U.S.Department of Health and Human Services. Common terminology criteria for adverse events (CTCAE) version 4.03.2010. USA: National Institutes of Health, National Cancer Institute. 2016.

45. Mittelstraß K, Waldenberger M. DNA methylation in human lipid metabolism and related diseases. Curr Opin Lipidol. 2018;29(2):116–24.

46. Wang Z, Wilson CL, Easton J, Thrasher A, Mulder H, Liu Q, et al. Genetic Risk for Subsequent Neoplasms Among Long-Term Survivors of Childhood Cancer. Journal of clinical oncology: official journal of the American Society of Clinical Oncology. 2018;36(20):2078–87.

47. Aryee MJ, Jaffe AE, Corrada-Bravo H, Ladd-Acosta C, Feinberg AP, Hansen KD, et al. Minfi: a flexible and comprehensive Bioconductor package for the analysis of Infinium DNA methylation microarrays. Bioinformatics. 2014;30(10):1363–9.

48. Du P, Zhang X, Huang CC, Jafari N, Kibbe WA, Hou L, et al. Comparison of Beta-value and M-value methods for quantifying methylation levels by microarray analysis. BMC Bioinformatics. 2010;11:587

49. Houseman EA, Accomando WP, Koestler DC, Christensen BC, Marsit CJ, Nelson HH, et al. DNA methylation arrays as surrogate measures of cell mixture distribution. BMC Bioinformatics. 2012;13:86.

50. Jaffe AE, Irizarry RA. Accounting for cellular heterogeneity is critical in epigenome-wide association studies. Genome biology. 2014;15(2):R31.

51. Barfield RT, Kilaru V, Smith AK, Conneely KN. CpGassoc: an R function for analysis of DNA methylation microarray data. Bioinformatics. 2012;28(9):1280–1.

52. Mansell G, Gorrie-Stone TJ, Bao Y, Kumari M, Schalkwyk LS, Mill J, et al. Guidance for DNA methylation studies: statistical insights from the Illumina EPIC array. BMC Genomics. 2019;20(1):366.

53. Yin L, Zhang H, Tang Z, Xu J, Yin D, Zhang Z, et al. rMVP: A Memory-efficient, Visualization-enhanced, and Parallel-accelerated tool for Genome-Wide Association Study. bioRxiv. 2020:2020.08.20.258491.

54. Chen H, Boutros PC. VennDiagram: a package for the generation of highly-customizable Venn and Euler diagrams in R. BMC Bioinformatics. 2011;12:35.

55. Benjamini Y, Hochberg Y. Controlling the False Discovery Rate: A Practical and Powerful Approach to Multiple Testing. Journal of the Royal Statistical Society Series B (Methodological). 1995;57(1):289–300.

56. Tingley D, Yamamoto T, Hirose K, Keele L, Imai K. mediation: R Package for Causal Mediation Analysis. Journal of Statistical Software. 2014;59(5):1–38.

57. Li M, Zou D, Li Z, Gao R, Sang J, Zhang Y, et al. EWAS Atlas: a curated knowledgebase of epigenome-wide association studies. Nucleic acids research. 2019;47(D1):D983–d8.

58. Aas PA, Otterlei M, Falnes PO, Vågbø CB, Skorpen F, Akbari M, et al. Human and bacterial oxidative demethylases repair alkylation damage in both RNA and DNA. Nature. 2003;421(6925):859–63.

59. Duncan T, Trewick SC, Koivisto P, Bates PA, Lindahl T, Sedgwick B. Reversal of DNA alkylation damage by two human dioxygenases. Proceedings of the National Academy of Sciences of the United States of America. 2002;99(26):16660–5.

60. Irvin MR, Zhi D, Joehanes R, Mendelson M, Aslibekyan S, Claas SA, et al. Epigenomewide association study of fasting blood lipids in the Genetics of Lipid-lowering Drugs and Diet Network study. Circulation. 2014;130(7):565–72.

61. Thaker VV, Esteves KM, Towne MC, Brownstein CA, James PM, Crowley L, et al. Whole exome sequencing identifies RAI1 mutation in a morbidly obese child diagnosed with ROHHAD syndrome. The Journal of clinical endocrinology and metabolism. 2015;100(5):1723–30.

62. Rohde K, Keller M, la Cour Poulsen L, Blüher M, Kovacs P, Böttcher Y.Genetics and epigenetics in obesity. Metabolism: clinical and experimental. 2019;92:37–50.

63. Spencer DM, Bilardi RA, Koch TH, Post GC, Nafie JW, Kimura K, et al. DNA repair inresponse to anthracycline-DNA adducts: a role for both homologous recombination and nucleotide excision repair. Mutation research. 2008;638(1-2):110–21.

64. Lieber MR. The mechanism of double-strand DNA break repair by the nonhomologous DNA end-joining pathway. Annu Rev Biochem. 2010;79:181–211.

65. Fu D, Calvo JA, Samson LD. Balancing repair and tolerance of DNA damage caused by alkylating agents. Nature reviews Cancer. 2012;12(2):104–20.

66. Ciccia A, Elledge SJ. The DNA damage response: making it safe to play with knives. Molecular cell. 2010;40(2):179–204.

67. Shibata A. Regulation of repair pathway choice at two-ended DNA double-strand breaks. Mutation research. 2017;803-805:51–5.

